# Native American Resilience to Protect Family Nutrition During a Pandemic: A Qualitative Analysis

**DOI:** 10.1101/2025.06.04.25329007

**Authors:** Sarah Vanegas, Reese Cuddy, Taylor Billey, Tanya Jones, Karlita Pablo, Novalene Goklish, Ashley Thacker, Leonela Nelson, Robin Tessay, Nicole Neault, Katie E. Nelson, Kimberlyn Yazzie, Allison Barlow

## Abstract

Native American (NA) communities have a history of being forced to adapt to adversity and leverage cultural strengths to cope with nutrition injustices. For generations, NA families in the United States (US) have experienced ongoing burdens from colonization-related disruptions to traditional foodways, resulting in disproportionately high rates of food and drinking water insecurity and related early childhood obesity. The COVID-19 pandemic further exacerbated these disparities. This study qualitatively explored NA families’ experiences with food access, water access and early childhood feeding during the COVID-19 pandemic. We further sought to identify resilience factors and specific response strategies that families employed to deal with challenges brought about by the pandemic. A total of 53 in-depth interviews were conducted with NA mothers of children 0-3 years old. Findings suggest that pandemic lockdowns and store restrictions magnified existing challenges and barriers to accessing food and water. Families leveraged resilience factors, including family support and practicing traditional foodways, to minimize negative impacts on child feeding practices. Understanding the social and cultural resilience factors used to cope with pandemic challenges from an Indigenous perspective can inform future strategies to improve food and water access and support positive child feeding practices for NA families, especially during public health crises.

## 1. Introduction

Pre-colonial Native American (NA) diets and food systems maintained healthy weight status and longevity among NA communities in the United States (U.S.). Presently, NA families experience significant nutrition inequities and associated health disparities stemming from a complex history of colonization and loss of lands that disrupted traditional food systems and lifeways (1,2). Compounding nutrition insecurity are social determinants including poverty, transportation barriers, a paucity of grocery stores, increasing food prices and water insecurity—all of which contribute to severe social, health and economic stress on families (3–5).

NA communities have had to demonstrate resilience in coping with nutrition injustices for generations. Resilience and other positive coping strategies are vital research constructs for understanding community-based assets to overcome health inequities. Understanding resilience from an Indigenous perspective offers an opportunity to promote wellbeing by identifying and leveraging cultural strengths to address root cause(s) of disparities (6).

Food insecurity, or the limited and uncertain availability of healthy food, disproportionately affects NA households in the U.S. Studies with Tribal communities have found household rates of food insecurity as high as 58%-92% in some regions (7–9) compared to 12.8% among all U.S. households (10). Food insecurity is a known risk factor for chronic disease, including obesity and type 2 diabetes which disproportionately affect NAs (11,12). NAs also face direct and disproportionate consequences from water insecurity (13,14). NA households are 19 times more likely to lack access to water and sanitation facilities compared to White households (15). Safe, adequate, and affordable access to water is essential to health, wellbeing, and nutrition. Disparities in NA water security stem from centuries of discrimination and neglect, colonization, forced removal and relocation, environmental racism, and long-standing underinvestment in essential water-related infrastructure across Tribal communities (16).

Early childhood feeding sets nutritional and weight status patterns for life. Responsive infant and toddler feeding practices positively impact healthy weight status across the lifespan (17). These practices include breastfeeding, developmentally appropriate introduction of solid foods, avoidance of sugar sweetened beverages, high intake of fruits and vegetables and low intake of processed snacks. Additionally, the way in which caregivers feed their children and model eating behaviors are as important to children’s healthy growth and weight status (18). Examples of healthy feeding methods include recognizing and responding appropriately to hunger and fullness cues and maintaining a division of feeding responsibilities between child and caregiver. However, ubiquitous food and water insecurity hamper caregiver’s capacity to establish ideal early childhood feeding patterns.

The COVID-19 pandemic has had an unprecedented impact on food insecurity, among low income, historically disenfranchised populations with pre-existing food insecurity (19). To our knowledge, only one study of COVID-related food has included a NA community in the U.S., the Blackfeet Nation, in which 79% of the study sample reported significantly greater food insecurity during the pandemic (20). Far less attention has been given to child feeding pattern changes and water insecurity resulting from the COVID-19 pandemic. The focus of this study was to identify impact of the COVID-19 pandemic on the experiences of rural NA families with young children related to child feeding patterns, food access, and water access. Due to the interrelatedness of these factors, we sought a holistic exploration, rather than examining any single factor. We also explored specific cultural strengths and resilience factors that caregivers employed from the direct perspectives of NA mothers. The specific questions we sought to answer were:

1. What challenges to food/water access and child feeding were experienced by NA households with young children during the COVID-19 pandemic?
2. What resilience factors enabled NA households to cope with these challenges?
3. What specific response strategies were employed by households to deal with these challenges?

This information can help shape future policies and programs that are responsive to the unique needs and strengths of Indigenous families, especially during times of crisis.

## 2. Methods

### 2.1 Study design

The current study is a qualitative analysis with a sub-sample of participants from the parent study, a randomized controlled trial of the Family Spirit Nurture program (22), which was disrupted due to the COVID-19 pandemic. The participating communities enacted strict policies to reduce the spread of the virus, which led both University IRB and Tribal IRBs to halt ongoing recruitment. Our team attempted remote delivery of the lessons, although this format affected participants’ engagement and retention in the intervention. In collaboration with onsite staff and our Data Safety Monitoring Board, we pivoted to explore updated study aims qualitatively based on resources and capacity.

### 2.2 Setting

Three reservation-based NA communities in the Southwestern U.S. participated: two communities on the Navajo Nation (Fort Defiance, AZ, and Shiprock, NM) and the White Mountain Apache community in Whiteriver, AZ. Food insecurity rates are exceedingly high on the Navajo Nation; in 2014, an estimated 76.6% of 175,000 Navajo Nation residents experienced some level of food insecurity (8). In 2022, there were only 14 grocery stores (23) across the Navajo Nation, roughly the size of West Virgina. Current estimates for water insecurity across Navajo Nation vary from 15-40% of the population; lack of reliable population-based data is a challenge (24–26). In the White Mountain Apache community, a population of 14,854 people, high rates of poverty (approximately 40%) and low rates of employment (37.7%) present significant challenges to food and water security (*Fort Apache Reservation, AZ - Census Bureau Profile*, 2022).

### 2.3 Research Team

The study leadership team consisted of senior level University researchers with PhD and MPH credentials, many with a decades-long history of working in partnership with Tribal communities. The data analysis team consisted of University Research Associates and an MPH graduate student. The community-based data collection team consisted of female, Indigenous, University Research Program Assistants, all of whom live and work in their respective Tribal communities and previously served as home visitors delivering the intervention and control curricula to study participants as part of the original parent study (22).

### 2.4 Ethics Approvals

This study was reviewed and approved by the Johns Hopkins University Bloomberg School of Public Health Institutional Review Board (7871) in June 2017. Additional approvals were received from the Navajo Nation Human Research Review Board (NNR-16.264), the Phoenix Area Indian Health Service IRB (PXR 17.05) in July, 2017.

### 2.5 Recruitment

We recruited a sub sample of 53 maternal participants from the 259 mother-child dyads that had been recruited for the parent study from across the three participating sites. The parent study specifically focused on the impact of maternal psychosocial factors on childhood outcomes; hence, the recruitment of a single-sex sample (22). Participants for this sub-study were personally invited either in person or via phone calls by community-based data collection team members to participate if they met the following inclusion criteria: consented participant in the parent study, at least 18 years of age, a NA parent/caregiver of child 0-3 years old. We aimed to recruit a sub sample representing 25% of mothers in both arms, chosen at random and stratified by demographics— food and water insecurity—a proxy for social economic status or challenging home conditions. With these strata in mind, we estimated that 25% of maternal participants from the parent study (approximately 18 participants from each of the three communities) would be more than sufficient to achieve data saturation (28). Of all 95 participants identified as eligible to complete interviews, 40 declined the invitation due to lack of time, discomfort talking about COVID-19, study staff unable to get a hold of participant, or no reason given by participant. Both currently enrolled participants and graduated participants were invited to participate. For those willing to participate, trained study staff obtained written informed consent for the interview. Recruitment for the original parent study began on October 16, 2017 and ended on March 17, 2020 (Covid Related). Recruitment for the qualitative interviews began on July 9, 2021 and ended on February 17, 2022.

### 2.6 Data collection

In-depth interviews (IDI) were conducted by study staff, all of whom are members of the participating Tribal communities and trained in qualitative research methods and participant privacy procedures. Our interview guide contained open-ended questions and prompts related to food and water access, child feeding patterns, and sugar sweetened beverage consumption among children. Questions were intended to capture experiences both before and during the COVID-19 pandemic. IDIs lasted between 30 and 90 minutes and were conducted in an agreed-upon, convenient location with participants (e.g., at the participant’s home, or the research program office, or via telephone), accounting for pandemic restrictions and participants’ preference. All participants were given a gift card upon completion of the interview. Interviews were digitally recorded, with participants’ consent, and transcribed verbatim (Rev.com). Participants were given the option to turn off the recording at any point during the interview if they felt uncomfortable. Field notes were made in REDCap software, a secure web-based application for managing databases, directly following interviews. The Supplemental Materials provide the full qualitative interview guide.

### 2.7 Data analysis

To begin, transcripts were reviewed for accuracy by study team members and uploaded to Atlas.ti (web version 2023), a qualitative data management software. We used a two-step, hybrid thematic analysis approach (29). First, 2 members of the study team developed a set of deductive codes based on our research questions to descriptively organize the data. Second, these coders inductively coded 3 randomly selected transcripts independently to explore the data in relation to relevant latent constructs and elicit key themes (29). They reached 80% agreement and met to discuss and resolve differences. The remaining interview transcripts were evenly divided between the 2 coders to complete the analysis. The analysis team met weekly during the coding process to discuss codes, iteratively refine the codebook, and prepare summary documents to share back with the Community Advisory Board, larger study team, and to each of the participating communities. Codebook available upon reasonable request.

## 3. Results

We conducted 53 in-depth IDIs with NA mothers of children 0-3 years of age. Demographic characteristics of participants are reported in **Table 1**. Three major themes were explored: 1) food and drinking water access challenges; 2) experiences of resilience; and 3) response strategies. In total, 80 sub-codes were identified and summarized to describe the range of experiences within each thematic area, which are visually represented in **Figure 1**. Quotations were chosen to represent the 3 participating communities equally. Quotations are included without participant’s age or community context due to Tribal Approval Board request that quotes remain anonymous and not traceable to individuals within the communities.

**Figure 1.**
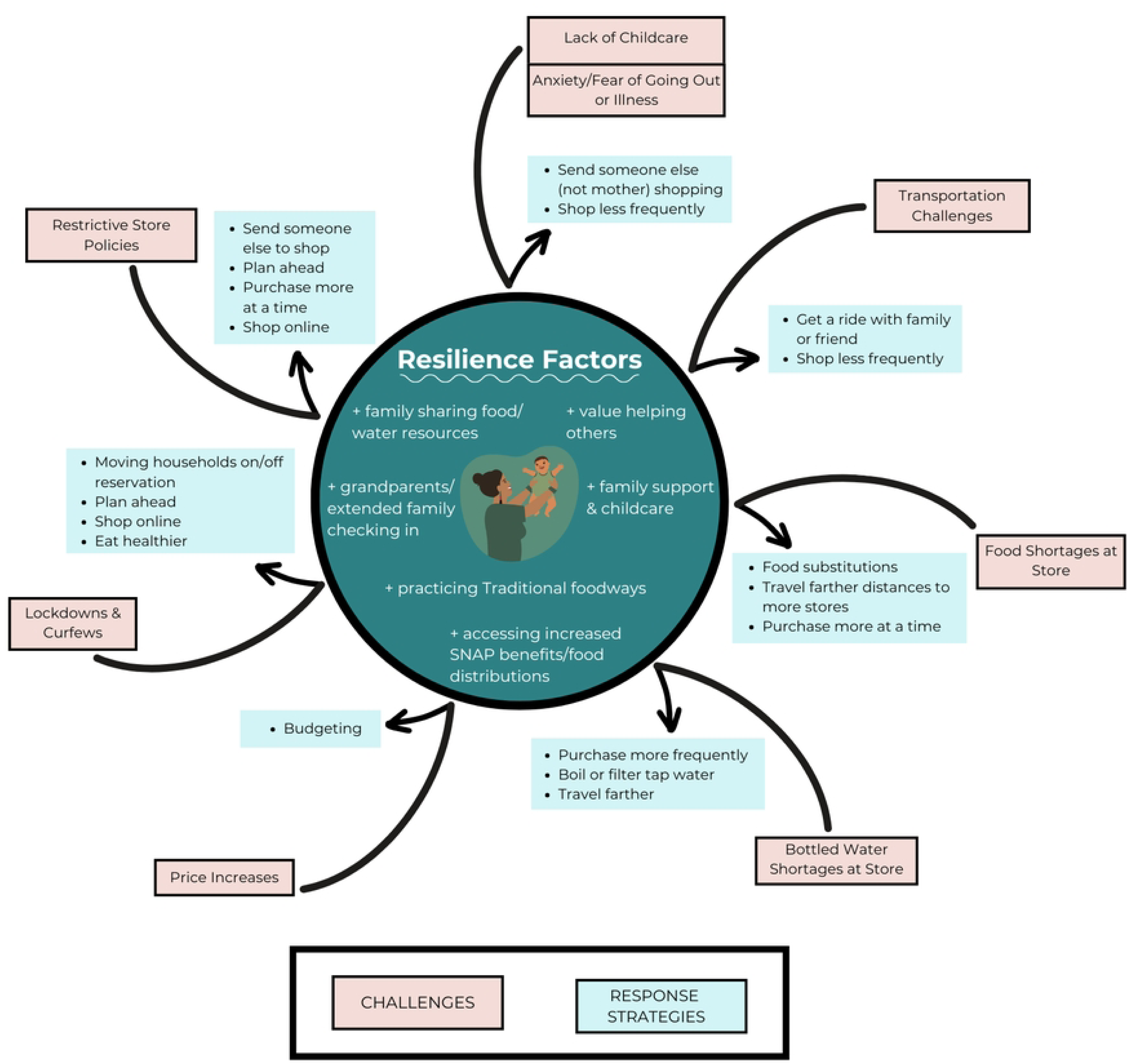
Resiliency framework illustrating the challenges and responses affecting food and water access during the pandemic. External challenges (shown in salmon colored boxes) related to the pandemic elicit response strategies (shown in turquoise boxes) shaped by Native American resilience factors (center circle).

**Table 1.** Participant demographics.

| Maternal baseline characteristics (N=53) | N (%) |
| --- | --- |
| **Communities:** |  |
| Fort Defiance, AZ (Navajo) | 18 (33.9%) |
| Shiprock, NM (Navajo) | 18 (33.9%) |
| Whiteriver, AZ (White Mountain Apache) | 17 (32.1%) |
| **Maternal age at enrollment: Mean (SD)** | 21.2 (2.3) |
| **Education** |  |
| Less than high school | 21 (39.6%) |
| Completed high school or GED | 18 (34.0%) |
| Completed > high School | 14 (26.4%) |
| **Married** |  |
| No | 47 (88.7%) |
| Yes | 6 (11.3%) |
| **Lived in > 1 home in the last year** |  |
| No | 32 (60.4%) |
| Yes | 21 (39.6%) |
| **Multigenerational household ($\geq 3$ generations) (N=52)** |  |
| No | 34 (65.4%) |
| Yes | 18 (34.6%) |
| **Food secure** |  |
| No | 18 (34.0%) |
| Yes | 35 (66.0%) |
| **Drinking Water secure** |  |
| No | 11 (20.8%) |
| Yes | 42 (79.2%) |
| **Employment status (N=53)** |  |
| No | 41 (77.4%) |
| Yes | 12 (22.6%) |
| Full-time employment | 5 (41.7%) |
| Part-time employment | 6 (50.0%) |
| Occasional | 1 (8.3%) |

| Over the past 12 months, generally, at the end of each month did you end up with |  |
| --- | --- |
| More than enough money left | 3 (5.7%) |
| Some money left over | 9 (17.0%) |
| Just enough to make ends meet | 25 (47.2%) |
| Almost enough to make ends meet | 8 (15.1%) |
| Not enough to make ends meet | 4 (7.5%) |
| Don't know | 4 (7.5%) |

### 3.1 Theme 1: Food and drinking water access challenges

Study participants shared the multitude of ways in which the COVID-19 pandemic impacted their ability to access food and drinking water—some new challenges, and others were exacerbated because of the pandemic. Beginning in March of 2020 when initial restrictions were implemented, families experienced food and water shortages, increased prices, social restrictions, anxiety, and other challenges.

Shortages of desired foods and bottled water at the grocery store became commonplace during the pandemic. Foods commonly mentioned as being hard to obtain were meat, infant formula, milk, water, and fresh produce. “*Hoarding*” or “*panic buying*” was commonly described as leading to these shortages. Families often drove to multiple stores trying to find essential items amid widespread shortages. In the Navajo Nation and White Mountain Apache community context, grocery stores are often located great distances apart or in neighboring communities 1-3 hours away. One participant explains, *“…when we got there, the whole shelves would be wiped out and, like, the meat would be all gone. So we had to go to another store and see if they have and if they didn’t have them, we gotta go to another store*.”

The increase in price of food and bottled water impacted many, especially those who experienced reduced or loss of work and income due to pandemic shutdowns. While some families noticed increased prices and were mindful of budgeting to make their money last, others did not notice any price increases due to the pandemic or it did not affect their ability to purchase what they needed. Some say they did not notice price increases because they were relying on others to shop for them and were not entering the store themself. Many families noted their increase in SNAP benefits allowed them to shop for what they needed without thinking about the price.

Mandatory lockdowns, curfews, and other restrictions put forth at the Tribal level inhibited families’ normal routines. For example, some communities required proof of residency to enter, which limited peoples’ ability to shop for groceries and other resources at certain hours, or in their usual locations. One mother shared, “*It was hard getting food during the lockdowns because I had both my kids and we had to go all the way to [a neighboring city] and get food and there was a lot of people there. So we all had to rush back for curfew.”*

Social restrictions along with anxiety related to fear of being exposed to the virus kept many participants from leaving their homes with their usual frequency. Strains on family relationships or the desire to be closer to family lead to many changes to living situations; participants describe moving in with other family members, separating and moving out, moving back onto the reservation, or moving off of the reservation during the pandemic. Multigenerational households both provided family support and presented challenges. For example, conflicting views on who was “*in charge*” or “*in control*” of food and drinks purchased and prepared for the family meals.

Procuring food and water became burdensome for parents with young children who needed to either find childcare or delegate grocery shopping to other family members. Additionally, for those without reliable transportation, lack of gas money or feeling uncertain about the acceptability of borrowing cars or asking for rides with others were described as daily issues. These new challenges contributed to grocery shopping happening less frequently. One mother described,

> *“It was kind of difficult because we had to travel farther, and at the time we had the old truck and it depended on how much money we had to put in for gas and who was going to watch baby for us. We couldn’t take her into the store. That’s where it kind of got difficult. … it just kind of slowed things down to where there was one person that had to go inside [the grocery store], and then after that, the next person would have to go inside. It would have to switch because the food was limited. It was just hard to get as much food but try to be very cautious on what was going on.”*

Ultimately, the pandemic forced families to become proactive about obtaining food and water, and those who did not, or were not able to, suffered the most. Those accessing food assistance programs like SNAP or WIC experienced challenges communicating with staff, had long wait times to get connected via phone, and providing eligibility documents to enroll was particularly cumbersome. One participant explained, “…*it was just too much to do. And then with our phone service, we had to update it… And once we finally got through with WIC … we had to wait for about an hour just to talk to somebody. But they wanted things to be faxed or mailed to them. We have nowhere to do that*.” Additionally, restricted store hours led many families to pivot to online grocery shopping. However, this type of access required planning ahead for pickup (i.e., child support, transportation, etc.) and reliable internet access to place orders.

### 3.2 Theme 2: Resilience Factors

Families described their ability to respond to many challenges due to factors like strong family support and practicing their Traditional foodways including self-reliant food procurement (i.e., hunting, fishing, growing produce). As depicted in **Figure 1**, these factors deflected pandemic-related challenges, and facilitated effective response strategies where needed.

First and foremost, participants shared stories of how strong family support networks and a value for helping others enabled them to get through difficult challenges brought on by the COVID-19 pandemic. The sudden inability to go to the store as needed, mandatory lockdowns and curfews, and changes to childcare access severely disrupted peoples’ lives. Many relied on family members for help with transportation, grocery shopping, and sharing means—e.g., money, food, and water—to get by. One participant shared, *“My mom was like a big help during the pandemic. She would… do stuff that I don’t think anybody else would’ve done. She would’ve like went to the store and all that… just to get like the food and all that for us…I guess you could say my mom is superwoman.”*

Changes to living situations, as a reaction to the pandemic, were described frequently. One mother who lived alone with her child before the pandemic and moved in with family members during the pandemic describes how everyone contributed to family wellbeing:

> *“…we did our best as a family to make sure [my child] was the safest and had everything she needed. So, if we had a struggle with like a utility bill or something, because everybody works in the family, um, certain people, let’s say my brother, would pitch in for that month on one utility each of the house, each of the people making an income would help with those things.”*

In conjunction with strong familial connections, practicing Traditional foodways enabled many families to be self-reliant and sustaining during a period of unprecedented food and water shortages nationwide. Multiple participants shared that they, themselves, or their extended family members engaged in traditional practices such as growing their own food, hunting, and fishing as a means for providing food for their families during the pandemic.

> *“We started growing our food because the pandemic and everything… We survived half of the time through that… That’s why we started having our own little garden and doing that and mixing vegetables with it and cooking with that. That also took a lot of the stress of trying to get food. That helped.”*

Flexibility, the ability to quickly adapt to changing situations, was a characteristic that some families described as aiding in their navigation of challenges. As one mother described, *“We’re kinda just, like, go with the flow kind of people. (laughs)….we decided if it works out this day [grocery shopping], then we’ll go, but if not, we’ll go this other day.”* This adaptability allowed families to access frequently shifting food resources during the pandemic. Increased SNAP benefits, and various local distributions of commodity foods were widely accessed and shared amongst family members.

### 3.3 Theme 3: Response strategies and child feeding pattern changes

Participants highlighted changes to their child(ren)s’ feeding patterns that directly resulted from the pandemic and how they responded. As noted above, store restrictions and childcare access often resulted in one parent having to stay home while the other retrieved food and necessary resources. In many instances, moms stayed home with the child(ren), which resulted in them having less control over what food and beverages were brought into the home and available to cook with. Those in multigenerational households sometimes noticed conflicting views regarding who should be “in charge” of what types of food to purchase and prepare for the household. One mother expressed,

> “[Before the pandemic] *I was the main cook, so I would always cook for my family. The one thing that changed was that I couldn’t really go in [to the grocery store] ‘cause I’m with the baby a lot. So the main person that would go is my mom or my boyfriend…and the one thing that mainly bugged me was that they didn’t get the food that I normally cook for my family. They would get like warmups and just something easy to make and not as tasty…I couldn’t really shop the way I wanted to.*”

Generally, participants reported that their child was getting the foods they wanted them to eat despite the pandemic. Despite low store availability of preferred food items, families were able to substitute other foods. For example, chicken or pork substituted for beef, 1% milk for whole milk, and frozen vegetables for fresh. Parents’ perceptions of healthy, desired foods for their children included vegetables, fruits, meats, and traditional foods such as blue corn mush and mutton. Where possible, some participants noted attempting to eat healthier, limiting junk/fried foods and cooking more meals at home, during the pandemic. One participant shared,

> *“[…] before the pandemic, we used to eat a lot of fast food, so we would go into town and eat a lot of hamburgers, pizza, chicken… And after the pandemic we’ve been keeping up with our nutrients. We try to cook at home all the time and try to eat a lot of vegetables, try to cook with a lot of vegetables.”*

Most families felt their child’s eating habits were generally healthy during the pandemic and reported little or no changes to the eating schedule or routine of their child due to pandemic impacts. Changes to eating habits during the pandemic were largely attributed to the child getting older and the resulting exposure to a wider range of foods and drinks (both healthy and unhealthy) and becoming more picky eaters with age.

Some participants became more aware of the importance of healthy nutrition principles during the pandemic. This resulted from receiving the Family Spirit Nurture lessons that were part of the parent study, having less time to peruse aisles in the grocery store resulting in decreased impulse buying of sugary foods and beverages, and increased mindfulness that accompanied less-busy schedules. Many families describe taking more time to enjoy being with family, being intentional about focusing on children, enjoying having partner/father of children around more often, talking with each other more, sharing mealtimes and creating routines and rituals as well as creating healthier eating habits. Conversely, some families describe eating less healthy during the pandemic due to increased snacking, stay-at-home boredom, or food preparers lack of motivation to prepare healthy meals, and reliance on microwaved or frozen meals.

To cope with shortages of bottled drinking water, a few participants mentioned boiling tap water, especially for use with mixing powdered infant formula or using a water filter. Tap water was generally undesirable for drinking because of taste and/or safety concerns. Though not common, one participant notably described being sickened by drinking contaminated water from a well with a high concentration of heavy metal. In response, she discontinued drinking it and increased her consumption of sugar sweetened beverages in an effort to save the limited amount of bottled water available for her young child.

Most families reported the COVID-19 pandemic did not significantly change the drinks that their child consumed; families were able to give their child the drinks they wanted to give them, which included water, milk, and small amounts of juice often diluted with water. Many respondents reported an increase in sugar sweetened beverages such as sodas and sports drinks due to the child getting older, changes in living situations, and family influences. As the child increased in age from infant to toddler, a wider range of drinks were available to them. Also, as a result of exposure to extended family members’ drinking sugary beverages and other family members offering soda to children, often against the mother’s wishes, children’s consumption of sugary drinks increased. One mother shared,*“I try my hardest to keep him away from it. But once he sees like a soda or like his grandma’s having a soda, he goes right for it.…So ever since his grandma has been giving him soda it’s like way more than he drank before”*

Families identified several areas of need for future desired community services. Transportation to grocery stores for all community members, more frequent distribution of food box delivery to households (rather than individual’s having to go to a location and wait in line), distribution of water filters to all households, community help hauling water, community services for Tribal elders, and assistance with lowering electricity bills were all mentioned as additional desired community services, especially during emergencies such as the COVID-19 pandemic.

> *“I think they could’ve offered, like, dropping off food at people’s doorstep instead of having to go out and, you know, waiting in line … with a lot of people there.”*
>
> *“I think at the beginning of the COVID, like when we didn’t really have water, I would think that it would’ve been better if the Chapter checked up on their community and helped haul water.”*
>
> *“…for my grandpa--he would walk to the store on essential days-and he had long ways to walk. But I couldn’t even go to help him shop because everybody had the time [store policy] for elderlies…I think it would’ve been, like, a big help if they checked on the elders more.”*

The inner circle of **Figure 1** represents the resilience factors identified in the IDIs - principally strong extended family support and a value for helping others. External pandemic challenges are deflected by the resilience factors and elicit the bulleted response impacts and strategies.

## 4. Discussion

The purpose of this study was to qualitatively examine the impacts of the COVID-19 pandemic on rural NA families’ ability to access food and water and maintain feeding patterns for their young children. Findings illustrate how specific resilience factors observed among Navajo and White Mountain Apache families deflected pandemic-related food and water access challenges to elicit protective response strategies. We heard clearly from participants that lockdowns and restrictive store policies presented new food and water access challenges and exacerbated pre-existing challenges for the three participating communities. In response to the challenges, families leveraged strong resilience factors, including family support and Traditional foodways, to minimize negative impacts on child feeding practices. Some positive and some negative changes to child feeding patterns were reported as a result of the pandemic, yet overall, families described the ability to maintain a regular eating schedule and routine for their children.

An important contribution of this study is that it demonstrates the strong resilience of NA families and their employment of specific social and cultural factors to cope with pandemic challenges. Supporting family, sharing resources, helping others, living in multigenerational households, practicing traditional foodways such as hunting and growing food, and being previously accustomed to traveling long distances for food and water are likely all factors that allowed families to report little to no changes in their child’s feeding patterns during the COVID-19 pandemic. These social resilience factors fit with the Indigenous Determinants of Health groundwork, increasingly used in research studies working in partnership with Tribal communities (30). It also contributes to the work of broadening the understanding of collective resilience from Indigenous perspectives and aligns with the notion that Indigenous people are strongest when they gather as community (6).

The main challenges to accessing food and water during the pandemic in all three study locations were broadly related to restrictive store policies, limited transportation, restricted mobility (e.g., lockdowns and curfews), price increases, and food and water shortages. These challenges align with the findings from previous studies focused on rural populations in high-income countries (31). The current study gathered additional insights in the context of rural, NA communities where individuals frequently live on reservation land and work in off-reservation communities, or otherwise maintain lives that straddle reservation borders. While the lockdowns of reservation communities and blockades to local roads and highways were enacted to protect communities and minimize the spread of the virus, these travel restrictions presented significant challenges for participants in this study including procuring food and water supplies and sharing those resources with family members—thereby inhibiting a core response strategy and source of resilience. The high incidence of reported changes to living situations and moving of households during the pandemic noted by study participants may be related to the lockdowns and travel restrictions and resource access challenges unique to these community contexts.

The importance of Indigenous community food sovereignty and robust local food systems are highlighted by pandemic restrictions. This is an increasingly fundamental area of work with the potential to minimize food shortages, price increases, and transportation and mobility challenges during everyday operations and global public health crises (32). It has also been stated that recognizing NA communities as food producers, and not solely food consumers, is an important part of the food decolonization process (33).

### 4.1 Strengths and limitations

Findings from the present study must be contextualized within its limitations. This study benefited from a relatively large sample size of participants (n=53) across three Tribal communities in the Southwest U.S. The themes described may share some commonalities to other communities due to the federal restrictions imposed during the pandemic but are nuanced given the broad regional differences among the 574 federally recognized Tribes across the U.S. Additionally, data were collected across a timespan that included various levels of ongoing COVID-19 pandemic restrictions, which may have influenced the interviewers’ ability to engage with participants. The rapidly changing landscape of the pandemic may have affected participants recall of COVID-19-related challenges and their overall ability to participate in the study. A key strength was that the interviewers were Indigenous researchers who live and work in the represented Navajo and White Mountain Apache communities. The study team, a mix of Indigenous and non-Indigenous researchers, were from an institution with a decades long history of working in partnership with Tribal communities to advance Indigenous well-being and health leadership to the highest level. This background of the study team was critical to the success of all aspects of the study.

### 4.2 Recommendations

Future studies could inform additional ways that resilience factors, especially from an Indigenous perspective, may protect from t+he impacts of future external crises. Given the current study’s theme of renewed interest in practicing Traditional foodways during a time of heightened food access barriers, more work is needed to augment local Native food systems. There is also a need for further exploration of how water insecurity impacts child feeding patterns, and how Indigenous water life ways tie to resilience.

## 5. Conclusion

The 53 NA mothers with young children who participated in this study’s qualitative interviews experienced heightened food insecurity and increased challenges with accessing food and water during COVID-19. They also described coping strategies and resilience factors that helped them to address the increased challenges and protect their family’s nutrition during a critical time of their young children’s development in a global crisis. In sharing experiences from a large sample of mothers’ food access, water access and child feeding practices in rural NA communities during COVID-19, our study adds to the literature that supports the importance of addressing food and water insecurity for NA communities, recognizing the protective factors of resilience from an Indigenous perspective, and furthering work in Indigenous food sovereignty. Especially as food insecurity rates among NA communities remain the highest in the US, and federal funding to nutrition programs may be unreliable, our recommendations focus on building sustainable local Native food systems that incorporate Traditional foodways based on the resilience factors and strengths-based coping strategies illustrated in this study’s interviews.

## Declaration Statements

### Author Contributions

Sarah Vanegas: Conceptualization, Methodology, Project administration, Data curation, Formal analysis, Writing – original draft, Writing – reviewing and editing

Reese Cuddy: Conceptualization, Methodology, Project administration, Data curation, Formal analysis, Writing – original draft, Writing – reviewing and editing Taylor Billey: Investigation, Data curation, Writing – reviewing and editing

Tanya Jones: Investigation, Data curation, Writing – reviewing and editing

Karlita Pablo: Investigation, Data curation, Writing – reviewing and editing

Novalene Goklish: Conceptualization, Methodology, Supervision, Writing – reviewing and editing Ashley Thacker: Project administration, Data curation, Formal analysis, Writing – reviewing and editing

Leonela Nelson: Investigation, Data curation, Writing – reviewing and editing

Robin Tessay: Investigation, Data curation, Writing – reviewing and editing

Nicole Neault: Conceptualization, Project administration, Formal analysis, Writing – original draft, Writing – reviewing and editing

Katie Nelson: Methodology, Writing – original draft, Writing – reviewing and editing

Kim Yazzie: Investigation, Data curation, Writing – reviewing and editing

Allison Barlow: Conceptualization, Methodology, Supervision, Writing – reviewing and editing

## Acknowledgements

We respectfully acknowledge the leadership, community members, and participants who guided, informed, and gave to this study from each of the participating communities.

## Data Availability

The data supporting this study’s findings are protected by Tribal sovereignty and are available pending Tribal approval upon reasonable request.

